# Alpha-synuclein seeds in olfactory mucosa of patients with isolated rapid-eye-movement sleep behaviour disorder

**DOI:** 10.1101/2020.08.04.20168328

**Authors:** Ambra Stefani, Alex Iranzo, Evi Holzknecht, Daniela Perra, Matilde Bongianni, Carles Gaig, Beatrice Heim, Monica Serradell, Luca Sacchetto, Alicia Garrido, Stefano Capaldi, Almudena Sánchez-Gómez, Maria Paola Cecchini, Sara Mariotto, Sergio Ferrari, Michele Fiorini, Joachim Schmutzhard, Pietro Cocchiara, Isabel Vilaseca, Lorenzo Brozzetti, Salvatore Monaco, M. Jose Marti, Klaus Seppi, Eduardo Tolosa, Joan Santamaria, Birgit Högl, Werner Poewe, Gianluigi Zanusso, for the SINBAR (Sleep Innsbruck Barcelona) group

## Abstract

Isolated REM sleep behaviour disorder is an early-stage α-synucleinopathy in most, if not all, affected subjects. Detection of pathological α-synuclein in peripheral tissues of isolated REM sleep behaviour disorder patients may identify those progressing to Parkinson’s disease, dementia with Lewy bodies or multiple system atrophy, with the ultimate goal of testing preventive therapies. <u>R</u>eal-<u>T</u>ime <u>Qu</u>aking-<u>I</u>nduced <u>C</u>onversion (RT-QuIC) provided evidence of α-synuclein seeding activity in cerebrospinal fluid and olfactory mucosa of patients with α-synucleinopathies. Aim of this study was to explore RT-QuIC detection of α-synuclein aggregates in olfactory mucosa of large cohort of subjects with isolated REM sleep behavior disorder compared to Parkinson’s disease and controls.

This cross-sectional case-control study was performed at the Medical University of Innsbruck, Austria, the Hospital Clinic de Barcelona, Spain, and the University of Verona, Italy. Olfactory mucosa samples obtained by nasal swab in 63 patients with isolated REM sleep behavior disorder, 41 matched Parkinson’s disease patients and 59 matched controls were analysed by α-synuclein RT-QuIC in a blinded fashion at the University of Verona, Italy. Median age of isolated REM sleep behavior disorder patients was 70 years, 85.7% were male. All participants were tested for smell, autonomic, cognitive and motor functions.

Olfactory mucosa was α-synuclein RT-QuIC positive in 44.4% isolated REM sleep behavior disorder patients, 46.3% Parkinson’s disease and 10.2% controls. While the sensitivity for isolated REM sleep behavior disorder plus Parkinson’s disease versus controls was 45.2%, specificity was high (89.8%). Among isolated REM sleep behavior disorder patients with positive α-synuclein RT-QuIC, 78.6% had olfactory dysfunction compared to 21.4% with negative α-synuclein RT-QuIC, p<0.001. The extent of olfactory dysfunction was more severe in positive than negative α-synuclein RT-QuIC olfactory mucosa isolated REM sleep behavior disorder patients (p<0.001).

We provide evidence that α-synuclein RT-QuIC assay enables the molecular detection of neuronal α-synuclein aggregates in olfactory mucosa of patients with isolated REM sleep behavior disorder and Parkinson’s disease. Although the overall sensitivity was moderate in this study, nasal swabbing is attractive as simple, non-invasive test and might be useful as part of a screening battery to identify subjects in the prodromal stages of α-synucleinopathies. Further studies are needed to enhance sensitivity, and better understand the temporal dynamics of α-synuclein seeding in the olfactory mucosa and spreading to other brain areas during the progression from isolated REM sleep behavior disorder to overt α-synucleinopathy, as well the impact of timing, disease subgroups and sampling technique on the overall sensitivity.

## Introduction

Isolated REM sleep behaviour disorder is characterized by abnormal behaviours during REM sleep (American Academy of Sleep Medicine, 2014). Long-term follow-up studies showed that more than 80% of patients with isolated REM sleep behaviour disorder may go on to develop Parkinson’s disease, dementia with Lewy bodies or, less commonly, multiple system atrophy (Schenck et al., 2013; Iranzo et al., 2013). These disorders are characterised by pathological deposition of α-synuclein (α-syn) aggregates in different sites within the central and peripheral nervous system and thus collectively labelled as α-synucleinopathies. Therefore, isolated REM sleep behaviour disorder is now commonly regarded as an early stage α-synucleinopathy (Högl et al., 2018).

Identification of early or prodromal stages of α-synucleinopathies is a key research goal on the path to disease-modifying and neuroprotective therapies. Recently, there has been great interest in detecting α-syn deposition in peripheral tissues of subjects with isolated REM sleep behaviour disorder as a potential biomarker of prodromal Lewy body disease stage. Intraneural phosphorylated α-syn (p-α-syn) deposits have been demonstrated by immunohistochemistry in tissue biopsies of colon (Sprenger et al., 2015), salivary glands (Vilas et al., 2016; Fernandez-Arcos et al., 2018; Iranzo et al., 2018) and skin of patients with Parkinson’s disease but also those with isolated REM sleep behaviour disorder (Antelmi et al., 2017; Doppler et al., 2017), where they were shown to differentiate isolated REM sleep behaviour disorder subjects from controls with variable sensitivity (24-89%) but overall high specificity (78-100%). However, these approaches are invasive, and feasibility and patient acceptance face limits when it comes to large scale screening for prodromal Parkinson’s disease or repeated prospective assessments of disease progression (Chanine et al., 2018). In addition, while colonic biopsies have tested the potential starting point of one suggested route of seeding and spreading of pathological α-syn species, i.e. from gut to brain, the olfactory system as the second proposed region of initiation of α-syn pathology in Parkinson’s disease has not been studied yet in prodromal Parkinson’s disease stages such as isolated REM sleep behaviour disorder (Ruffmann et al., 2018). <u>R</u>eal-<u>T</u>ime <u>Qu</u>aking <u>I</u>nduced <u>C</u>onversion (RT-QuIC) is a novel assay based on the so-called “prion replication principle” implying that pathologic misfolded proteins (seeds) serve as template for imparting their conformation to normal isoform (substrate). Tissue samples such as cerebrospinal fluid and olfactory mucosa (OM) containing α-syn aggregates initiate amyloid fibril formation by converting the recombinant α-syn which, in turn, enhances the fluorescence of thioflavin T (ThT) (Fairfoul et al., 2016; De Luca et al., 2019; Garrido et al., 2019).

Olfactory dysfunction is common in patients with isolated REM sleep behaviour disorder, where it represents a predictor of short-term phenoconversion to Parkinson’s disease or dementia with Lewy bodies (Mahlknecht et al., 2015), and is almost universal in established Parkinson’s disease. The olfactory dysfunction in Parkinson’s disease is likely related to Lewy pathology and neuronal cell loss in the olfactory bulbs, tracts and piriform cortex, and according to the Braak staging the olfactory bulbs may be an initial site of α-syn aggregation in Parkinson’s disease (Braak et al., 2003; Rey et al., 2018). The misfolded α-synuclein, which replicates and propagates with a prion-like mechanism, is believed to drive the neurodegenerative process.

In the present study, we tested a novel biomarker approach for identifying α-syn aggregates by RT-QuIC assay in OM samples obtained from patients with isolated REM sleep behaviour disorder, established Parkinson’s disease and healthy controls.

## Materials and methods

### Study design and participants

This was a cross-sectional study performed at three clinical academic centers (Department of Neurology of Innsbruck Medical University, Austria; Hospital Clinic de Barcelona, Spain; Neurology Clinic, Department of Neurosciences, Biomedicine and Movement Sciences of the University of Verona, Italy). The study was approved by the local ethics committees and all participants provided written informed consent according to the [Declaration of Helsinki](https://www.wma.net/policies-post/wma-declaration-of-helsinki-ethical-principles-for-medical-research-involving-human-subjects/).

Sixty-three patients with polysomnography-confirmed isolated REM sleep behaviour disorder were recruited at the Sleep Disorder Units of the Department of Neurology, Innsbruck Medical University, Austria, and Hospital Clinic de Barcelona, Spain. Isolated REM sleep behaviour disorder was diagnosed according to the current International Classification of Sleep Disorders criteria (American Academy of Sleep Medicine, 2014), in the absence of parkinsonism or cognitive impairment (excluded by clinical history and examination). Demographic and clinical data were collected through interview and review of medical records.

The Movement Disorder Units of Innsbruck and Barcelona clinical departments and the Neurology Clinic in Verona recruited age- and sex-matched Parkinson’s disease patients (n=41), diagnosed according to the Movement Disorders Society (MDS) clinical diagnostic criteria (Postuma et al., 2015). Fifty-nine age- and sex-matched controls without parkinsonism or cognitive impairment were also included. Controls were recruited in Innsbruck from patients of the sleep laboratory in whom isolated REM sleep behaviour disorder and REM sleep without atonia were excluded by video-polysomnography (performed no longer than six months before OM sampling), and in Barcelona from non-blood relatives of isolated REM sleep behaviour disorder or Parkinson’s disease patients participating in the study. In all controls, evidence for a neurodegenerative disease was excluded by clinical interview and neurological examination. For control subjects without availability of a video-polysomnography, dream-enacting behaviours were excluded by clinical history.

### Clinical markers of α-synuclein related neurodegeneration

In all participants, smell function was assessed with the 16-item identification part of the sniffin’sticks (Oleszkiewicz et al., 2019) or with the 40-item University of Pennsylvania Smell Identification Test (UPSIT) (Doty, 1995), the presence and quantification of motor signs using the motor part of the MDS Unified Parkinson’s Disease Rating Scale (MDS-UPDRS) (Goetz et al., 2008), autonomic dysfunction with the SCOPA (SCales for Outcomes in PArkinson’s disease)-AUT (autonomic dysfunction) (Visser et al., 2004), and cognitive function with the Montreal cognitive assessment (MoCA) (Nasreddine et al., 2005). These evaluations were performed no longer than three months before OM sampling. Additionally, in Parkinson’s disease patients a history suggestive of isolated REM sleep behaviour disorder was obtained using the REM sleep behaviour disorder (RBD) single question screen (RBD1Q) at the day of OM sampling (Postuma et al., 2012).

For data analysis, UPSIT scores were converted to Sniffin’ 16 scores using published equating scores (Lawton et al., 2016). Olfactory dysfunction was defined according to published updated age- and sex-adjusted normative data for the Sniffin’ 16 scores, using the 10^th^ percentile as cut-off (Oleszkiewicz et al., 2019).

MDS prodromal PD calculation was performed for iRBD patients, according to the updated MDS research criteria for prodromal PD (Heinzel et al., 2019), using the online prodromal PD calculator (http://www.movementdisorders.org/MDS/Members-Only/Prodromal-PD-Calculator.htm). For patients aged < 50 years we selected the age range 50-54 years (the lowest age range provided by the calculator).

### Olfactory mucosa sampling

Nasal swabbing procedure was performed in all participants using specifically designed flocked brush (FLOQBrush®; CopanItalia Spa, Brescia, Italy), as described previously (Bongianni et al., 2017). OM sampling was performed by otolaryngologists (LS, JS and IV) and did not require local anesthesia. The nasal swabbing procedure took less than five minutes and was done without the use of nasal tampons, to avoid patients’ discomfort or airways obstruction (see video 1). Coagulation disorder or anticoagulant/antiplatelet drug intake or other medical conditions were not an exclusion criterion. Adverse events were immediately recorded. At the end of the procedure, 61 participants (37.4%) were asked for evaluating the degree of pain or discomfort perceived during nasal swabbing on a scale from one (i.e., minimal discomfort) to 10 (i.e., maximal discomfort).

One to four OM samples were collected from each individual, depending on nasal cavity anatomy and individual tolerability. Almost all subjects had two nasal swabbings taken from a single nostril, except twelve subjects in whom sampling was performed bilaterally. Since two swabs were required for RT-QuIC analysis cytological quality control of samples was limited to the twelve subjects with bilateral procedures. Eight of these samples were processed for immunocytochemical analysis and from a single swab we collected one million of total cells where 30% were olfactory marker protein positive (Brozzetti et al., 2020).

### Olfactory mucosa sample preparation

Following nasal swabbing, the swab was immersed in a 5 ml tube containing 0.9% saline and sealed. Tubes from each patient were labelled with an anonymised code, stored at 4°C, and sent to Neuropathology laboratory at University of Verona, Italy. Cellular material was dissociated from the swab by vortexing tubes for 1 min at room temperature. Then, the swab was removed from the tube, and cell suspension was pelleted by centrifugation at 2000×*g* at 4°C for 20 minutes. The supernatant was removed, and pellet frozen at −80°C until assayed.

### Alpha-synuclein RT-QuIC analysis in olfactory mucosa swabs

RT-QuIC analyses were performed in the LURM (Laboratorio Universitario di Ricerca Medica) Research Center, University of Verona, Italy. Recombinant α-syn was expressed and purified from the periplasmic fraction as previously reported (Bongianni et al., 2019). The α-syn RT-QuIC test used in this study has been previously setup using brain tissue of definite cases of Parkinson’s disease, dementia with Lewy bodies, and multiple system atrophy (Bongianni et al., 2019). OM samples were thawed and a disposable inoculating loop (Fisherbrand) was dipped into the pellet to transfer approximately 2 μl of the pellet into a tube containing 120 μl PBS. The latter tube was sonicated at 120W (Digital ultrasonic bath Mod.DU-32, Argo Lab) for at least one minute until the pellet was dispersed. For each test, two microliters of diluted OM sample were plated in 98 µl of Reaction Buffer composed of 100 mmol/L phosphate buffer (pH 8.2), 10 *μ*mol/L ThT, and 0.05 mg/mL human recombinant full length (1–140aa) α-syn and mg of 0.5-mm glass beads (Sigma). The plate was sealed with a plate sealer film (Nalgene Nunc International) and then incubated at 30°C in a BMG FLUOstar Omega plate reader with cycles of 1 min shaking (200 rpm double orbital) and 14 min rest. ThT fluorescence measurements (450+/-10 nm excitation and 480+/-10 nm emission; bottom read) were taken every 45 min. Four replicate reactions were tested for each sample. ThT fluorescence threshold was calculated considering three SD above the baseline. Because of the initial increase of fluorescence signal in all curves, the baseline was determined between 15 and 17 hours.

A sample was considered positive when at least two out of four replicate wells crossed this calculated threshold (100,000 rfu). If only one of the replicates was positive, the RT-QuIC was considered to be negative. Cut-off time was assessed at 80 hours, based on the results from definite cases, in order to obtain the best specificity and sensitivity.^29^ The maximal fluorescence value was the highest mean fluorescent value seen during the α-syn RT-QuIC analytical run of 80 h and we considered the lag-phase as the hours required for the average fluorescence to exceed the threshold for individual cases, as reported in figure 1. At the end of sample analyses, RT-QuIC results were sent to the study coordinator (AS) who unblinded the results.

**Figure 1.**
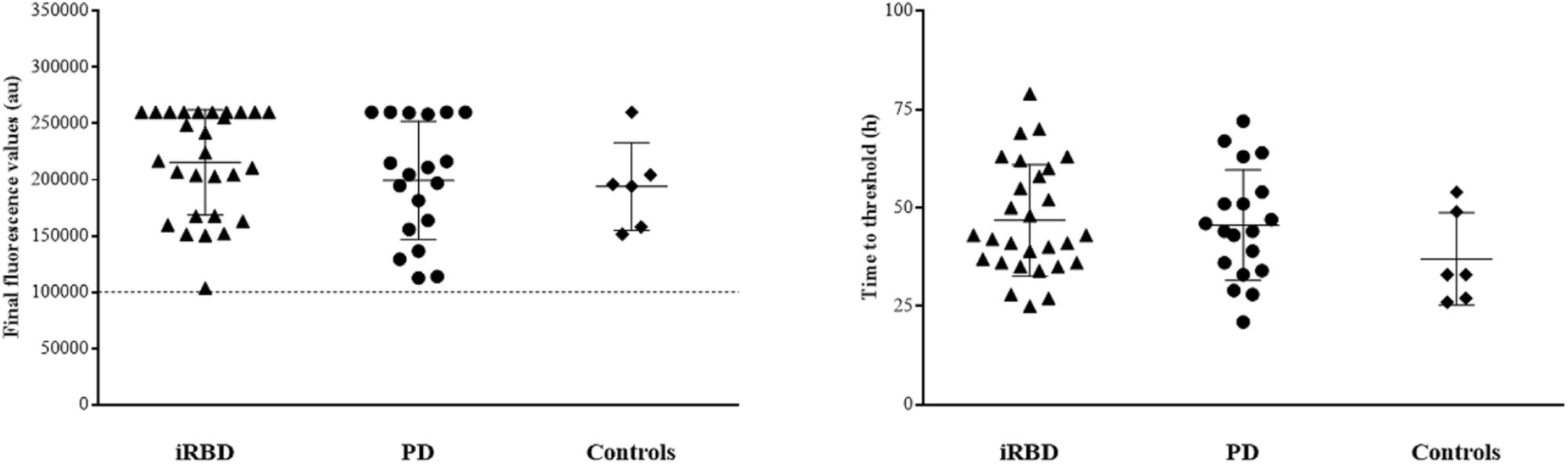
Real-Time Quaking-Induced Conversion (RT-QuIC) analysis of OM samples from patients with isolated RBD, Parkinson’s disease and controls. Final fluorescence values (a) and lag-phase (b) of positive α-syn RT-QuIC subjects in OM samples from iRBD, PD and controls. Data points in panel a, represent the average fluorescence value obtained for each individual case at 80h; samples are grouped in 3 different classes (iRBD, triangle; PD, circle; Controls, diamond) and bars show the average ± SD for type of case. Data points in panel b show hours required from the average fluorescence value to exceed the threshold of 100,000 rfu for individual cases; samples are grouped in 3 different classes (iRBD, triangle; PD, circle; Controls, diamond) and bars show the average ± SD for type of pathology. Au, arbitrary unit; h, hours; iRBD, isolated REM sleep behaviour disorder; PD, Parkinson’s disease.

### Statistical analysis

Data distribution was tested using the Kolmogorov-Smirnov or the Shapiro Wilk test. Since data were not normally distributed, quantitative variables are reported as median (interquartile range, IQR) and qualitative variables as number (%). Quantitative variables were analysed using the Kruskall-Wallis test for overall comparisons between groups. Qualitative variables were analysed with the Fisher’s exact test in case of comparison of two variables with two categories, with the Chi-squared test in case of two variables with more than two categories.

Sensitivity and specificity for OM+ in isolated REM sleep behaviour disorder plus Parkinson’s disease were calculated. Correlations were evaluated using the Spearman-Rho, Pearson correlation coefficient or Phi correlation coefficient, as appropriate. All statistical analyses were performed with SPSS (IBM SPSS Statistics, Version 25) and with STATA/IC 16.0 for Windows (StataCorp LLC). P values <0.05 were considered statistically significant.

## Data availability

Data can be available upon request to the corresponding author.

## Results

This study was performed between 2017 and 2020. A total of 163 participants were included: 63 patients with isolated REM sleep behaviour disorder, 41 Parkinson’s disease and 59 controls. Demographic characteristics and results of clinical assessments are summarized in Table 1.

**Table 1.** Demographic and clinical characteristics of the study population.

|  | **iRBD (n=63)** | **PD (n=41)** | **Controls (n=59)** | **P value** |
| --- | --- | --- | --- | --- |
| **Age, years** | 70 (64-74) | 70 (65-76) | 70 (64-73) | 0.690 |
| **Sex, n (%)** |  |  |  | 0.735 |
| **Female** | 9 (14.3) | 8 (19.5) | 11 (18.6) |  |
| **Male** | 54 (85.7) | 33 (80.5) | 48 (81.4) |  |
| **Disease duration (from diagnosis), y** | 5 (2-9) | 6.5 (2-9.8) | n.a. | n.a. |
| **MDS-UPDRS III score** | 4 (2-7) | 21 (13-30) | 1 (0-3) | **<0.001**<br>**iRBD vs PD <0.001**<br>**iRBD vs Ctr <0.001**<br>**PD vs Ctr <0.001** |
| **SCOPA-AUT score** | 14 (9-27) | 16 (9-23) | 11 (6-19) | 0.052<br>iRBD vs PD 0.724<br>**iRBD vs Ctr 0.023**<br>PD vs Ctr 0.075 |
| **MoCA score** | 27 (24-28) | 27 (24-28) | 27 (25-29) | 0.833<br>iRBD vs PD 0.793<br>iRBD vs Ctr 0.526<br>PD vs Ctr 0.846 |
| **Sniffin'sticks score** | 8 (7-12) | 8 (6-10) | 12 (11-13) | **<0.001**<br>iRBD vs PD 0.310<br>**iRBD vs Ctr <0.001**<br>**PD vs Ctr <0.001** |
| **Olfactory dysfunction, n (%)** | 30 (47.6) | 25 (61) | 8 (13.6) | **<0.001**<br>iRBD vs PD 0.229<br>**iRBD vs Ctr <0.001**<br>**PD vs Ctr <0.001** |

Data are shown as median (IQR) or N (%). Ctr, controls. iRBD: isolated REM sleep behaviour disorder; MDS-UPDRS III: Movement Disorders Society Unified Parkinson's Disease Rating Scale, part III; PD: Parkinson's disease; SCOPA-AUT: Scales for Outcomes in Parkinson's disease AUTonomic dysfunction; MoCA: Montreal cognitive assessment.

Olfactory function was similarly impaired in both isolated REM sleep behaviour disorder and Parkinson’s disease as compared to controls (table 1).

According to the MDS research criteria for prodromal Parkinson’s disease, 41/63 patients with isolated REM sleep behavior disorder (65.1%) met criteria for probable prodromal Parkinson’s disease (median estimated probability 98.3%, IQR 69.5%-99.9%). When excluding PSG-proven REM sleep behavior disorder from the calculation, 18/63 (28.6%) were still classified as probable prodromal Parkinson’s disease (median estimated probability 32.1%, IQR 1.4%-90.3%).

### OM sampling and α-synuclein RT-QuIC assay

Two nasal swabbings were performed into the most easily accessible nostril: on the left side in 102 (62.6%), on the right side in 49 (30.1%) participants, and bilaterally (two from each nostril because both were easily accessible) in 12 (7.4%). Nasal swabbing was not associated with adverse events or complications except for a short and transient local discomfort during the procedure. The latter was experienced as a sudden and brief (seconds) intense discomfort (score ≥ eight) by 9.8%, moderate discomfort (score five to seven) by 42.6%, only mild discomfort (score < five) by 47.6%.

Nasal swabs were RT-QuIC positive for α-syn in 28/63 (44.4%) patients with isolated REM sleep behaviour disorder, in 19/41 (46.3%) Parkinson’s disease and in six out of 59 (10.2%) controls (figure 1). RT-QuIC positivity for α-syn was significantly different between isolated REM sleep behaviour disorder and controls (p<0.001), as well as between Parkinson’s disease and controls (p<0.001), but not between isolated REM sleep behaviour disorder and Parkinson’s disease (p=0.504). Sensitivity for isolated REM sleep behaviour disorder and Parkinson’s disease versus controls was 45.2% (95% CI 35.4-55.3%), specificity 89.8% (95% CI 79.2-96.2%).

There was no significant difference in RT-QuIC responses, either of average final ThT fluorescence value (214720±48471 and 199415±52560 rfu) or lag-time phase (47±14.2 and 45±14 hours) between OM samples from patients with isolated REM sleep behaviour disorder and with Parkinson’s disease. Six of 59 OM samples from controls were positive within 80 hours of seeding reaction (figure 1, table S1).

### RT-QuIC assay results and clinical data

Twenty-two (78.6%) of the 28 OM-positive patients with isolated REM sleep behaviour disorder had olfactory dysfunction, while among the 35 OM-negative patients with isolated REM sleep behaviour disorder olfactory dysfunction was present in 22.9% (eight out of 35). The extent of olfactory dysfunction in isolated REM sleep behaviour disorder OM-positive patients was more severe compared to OM-negative subjects (p<0.001); moreover, there was a moderate correlation between RT-QuIC results and smell test score (Pearson correlation coefficient −0.490, p<0.001; after adjustment for age Pearson correlation coefficient −0.429, p<0.002; after adjustment for sex Pearson correlation coefficient −0.500, p<0.001) and a strong relationship with olfactory dysfunction (Phi coefficient 0.554, p<0.001).

These associations were not found in patients with Parkinson’s disease (table 2). Smell test results for the different study groups in relation to RT-QuIC results are shown in figure 2.

**Table 2.** Demographic and clinical characteristics of the iRBD group (n=63) and of the PD group (N=41).

|  | iRBD |  |  | PD |  |  |
| --- | --- | --- | --- | --- | --- | --- |
| | α-syn positive<br>n=28 (44.4%) | α-syn<br>negative<br>n=35 (55.6%) | P value | α-syn positive<br>n=19 (46.3%) | α-syn<br>negative<br>n=22 (53.7%) | P value |
| Age, y | 72 (67-74.8) | 67 (60-74) | 0.059 | 70 (65-75) | 71 (63.8-76.3) | 0.704 |
| Age at<br>diagnosis, y | 67 (63-68.8) | 60 (56-67) | **0.004** | 64 (57-68) | 64 (58-71) | 0.708 |
| Age at onset, y | 62 (59-67.5) | 53 (45-61) | **0.011** | 64 (56-68) | 63 (57-69) | 0.936 |
| Disease<br>duration, y | 4.5 (1.3-8) | 6 (2-10) | 0.176 | 7 (1-12) | 5 (2-9) | 0.934 |
| Sex, n (%) |  |  | 0.170 |  |  | 0.703 |
| Female | 6 (21.4) | 3 (8.6) |  | 3 (15.8) | 5 (22.7) |  |
| Male | 22 (78.6) | 32 (91.4) |  | 16 (84.2) | 17 (77.3) |  |
| MDS-UPDRS<br>III score | 4 (1-7) | 4 (2-7) | 0.692 | 21 (13-30) | 21 (12-28) | 0.937 |
| SCOPA-AUT<br>score | 18 (11-29) | 13 (8-21) | 0.133 | 15 (10-20) | 19 (8-26) | 0.432 |
| MoCA score | 27 (24-28) | 27 (24-28) | 0.737 | 27 (23-28) | 27 (24-28) | 0.926 |
| Sniffin'sticks<br>score | 7 (5-8) | 11 (8-13) | **<0.001** | 7 (6-11) | 8 (7-9) | 0.968 |
| Olfactory<br>dysfunction, n<br>(%) | 22 (78.6%) | 8 (22.9%) | **<0.001** | 11 (57.9%) | 8 (36.4%) | 0.757 |
| Hoehn and<br>Yahr stage | n.a. | n.a. | n.a. | 2 (2-2) | 2 (2-2) | n.a. |
| Rigidity, n (%) | n.a. | n.a. | n.a. | 13 (100%) | 21 (95.5%) | 1.000 |
| Rest tremor, n<br>(%) | n.a. | n.a. | n.a. | 7 (53.8%) | 12 (54.5%) | 0.536 |

Data are shown as median (IQR) or N (%). iRBD: isolated rapid-eye-movement sleep behaviour disorder; MDS-UPDRS III: Movement Disorders Society Unified Parkinson's Disease Rating Scale, part III; PD, Parkinson's disease; RBD1Q: REM sleep behaviour disorder single question; SCOPA-AUT: Scales for Outcomes in Parkinson's disease AUTonomic dysfunction; MoCA: Montreal cognitive assessment. P values were calculated using the Mann-Whitney U test.

**Figure 2.**
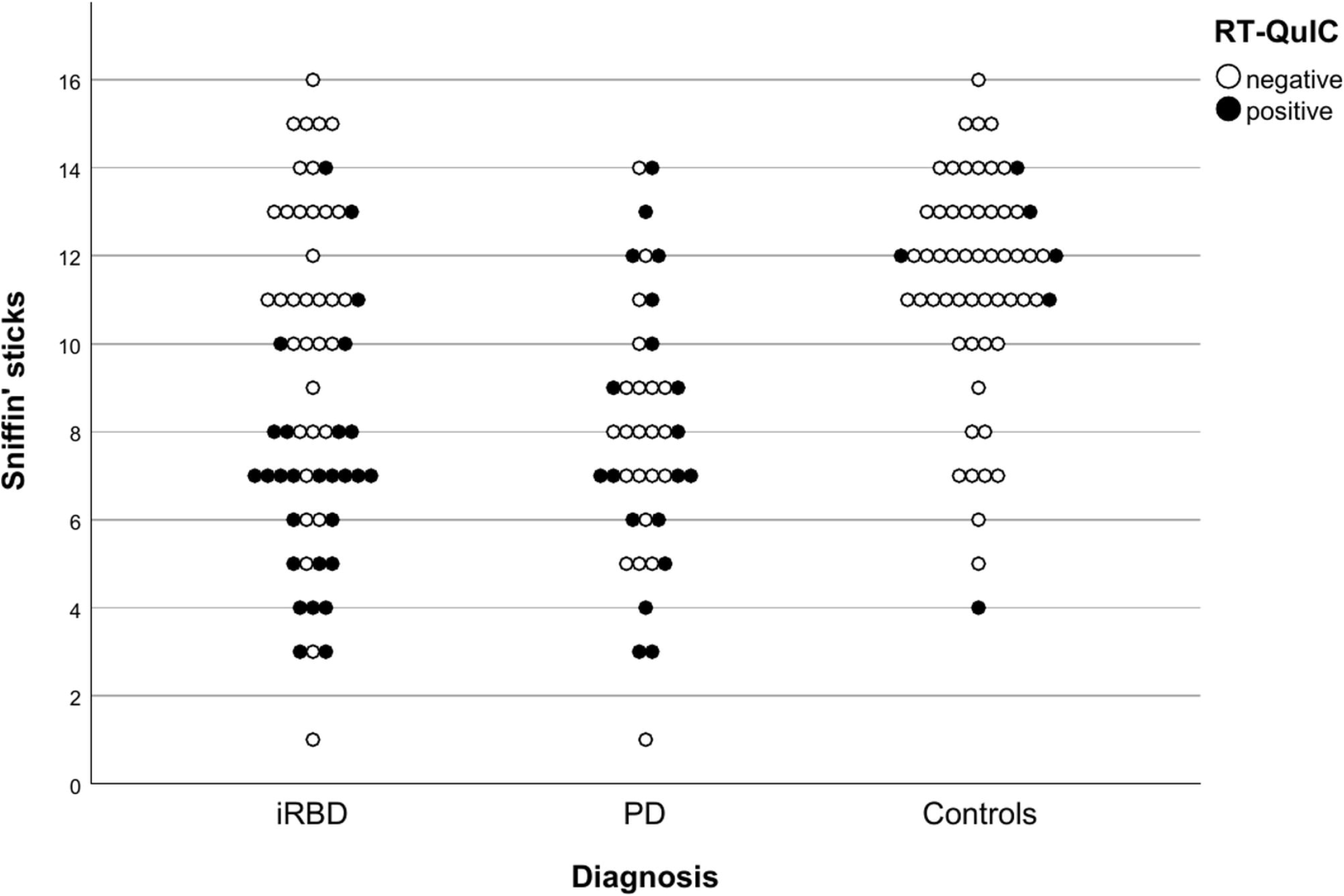
Olfactory mucosa (OM) Real-Time Quaking-Induced Conversion (RT-QuIC) for α-synuclein results and smell test scores in iRBD, PD and controls groups. Positive OM results are shown as a black circle, negative OM results are shown as a white circle. iRBD, isolated REM sleep behaviour disorder; PD, Parkinson’s disease; RT-QuIC, real-Time Quaking-Induced Conversion.

The median estimated probability for prodromal Parkinson’s disease based on the MDS criteria (excluding polysomnographic proven REM sleep behavior disorder from the calculator) was numerically higher in OM-positive versus OM-negative subgroup (median estimated probability 54.4%, vs 14.8%) but the ranges were large and the difference was not statistically significant (IQR’s 1.5%-72.2% and 1.2%-89.9%, p=0.782). Likewise, the percentages of patients meeting the threshold for a diagnosis of probable prodromal Parkinson’s disease were not statistically different between the two groups (17.9% vs 34.3%, p=0.166).

Among patients with Parkinson’s disease, 28/41 (68.3%) screened positively for REM sleep behaviour disorder with the RBD1Q. Positive response to the RBD1Q did not differ between OM-positive and OM-negative Parkinson’s disease patients (12/19, 63.2% vs. 16/22, 72.7%, p=0.737). We did not find any difference between PD with and without probable RBD regarding OM-positivity and sniffin’ stick score (Pearson correlation 0.129, p=0.676 in PD without probable RBD; Pearson correlation 0.007, p=0.972 in PD with probable RBD). The same was true for the correlation between olfactory dysfunction and OM-positivity (Phi coefficient = −0.141, p=0.612 in PD without probable RBD, Phi coefficient = −0.062, p= 0.743 in PD with probable RBD).

Further demographic and clinical characteristics of the isolated REM sleep behaviour disorder and Parkinson’s disease patients did not differ between subjects with or without OM α-syn seeding activity (table 2).

Among healthy controls, the six (10.2%) OM-positive individuals as a group did not differ from OM-negative controls in any of the assessed clinical parameters.

## Discussion

To the best of our knowledge this is the first study evaluating α-syn RT-QuIC in the OM of a large cohort of patients with isolated REM sleep behaviour disorder versus those with Parkinson’s disease or healthy controls. We studied a total of 163 subjects and found a positive α-syn RT-QuIC seeding reaction in the OM in 44.4% of patients with isolated REM sleep behaviour disorder, in 46.3% of those with Parkinson’s disease and also in 10.2% of controls. From the perspective of diagnostic testing this would result in an overall specificity of 89.8%, sensitivity of 45.2% and accuracy of 61.3%. Such figures, although impressive in terms of specificity, do not seem superior to what has been reported for immunohistochemical assays for p-α-syn in salivary glands and skin biopsies (Sprenger et al., 2015; Vilas et al., 2016; Antelmi et al., 2017; Doppler et al., 2017; Fernandez-Arcos et al., 2018; Iranzo et al., 2018). However, there are distinctive and unique features in the method employed in the present study.

OM sampling provides direct access to olfactory neurons and thus to one of the sites, that is currently regarded as one of the potential initiation points of seeding and spread of pathological α-syn assemblies in Parkinson’s disease. RT-QuIC for α-syn detects disease-associated α-syn aggregates in billion-fold diluted brain tissue preparations from different α-synucleinopathies (Groveman et al., 2018) with a higher sensitivity than conventional immunohistochemistry, as shown in prion diseases (Race et al., 2019). RT-QuIC results are classified as negative or positive based on the extent of α-syn aggregates seeding and do not depend on observer evaluation, as occurs for α-syn immunohistochemistry in skin biopsies of patients with iRBD (Donadio et al., 2019).

Furthermore, immunohistochemistry is too insensitive to detect the early misfolded forms of α-syn occurring in olfactory neurons, as suggested by the low yield of Lewy pathology in olfactory neuroepithelium of subjects with α-synucleinopathies (Arnold et al., 2010). In addition, RT-QuIC positivity for α-syn has been found in OM of 10% of controls, suggesting that α-syn aggregation occurs as an incidental event in the olfactory neuroepithelium (Saito et al., 2016). However, this finding was not unexpected since the occurrence of incidental Lewy bodies is around 10% in people over 60 years (Gibb et al., 1988; Beach et al., 2009) which corresponds to the mean age of controls recruited in the present study. It is intriguing to speculate if these individuals might be at increased risk to develop Parkinson’s disease or other α-synucleinopathies, but numbers here were too small for cross-sectional analysis.

In the only previous study (De Luca et al., 2019) using α-syn RT-QuIC on OM samples from patients with synucleinopathies (18 Parkinson’s disease, 11 multiple system atrophy vs 18 controls with non α-syn related disorders) sensitivity and specificity were broadly similar (56% and 83%, respectively) to the present study, supporting the reproducibility of the analytical approach.

Recent studies have also explored the performance and diagnostic accuracy of α-syn RT-QuIC assay of CSF in patients with α-synucleinopathies. In patients with Parkinson’s disease, diagnostic sensitivity and specificity were 84% and 89% respectively regarding differentiation from non-synucleinopathy parkinsonism (van Rumund et al., 2019).

Since the olfactory system is believed to be one of the earliest sites of pathology in PD, RT-QuIC for α-syn in OM samples is a particularly attractive approach not only in the quest for biomarkers of α-synucleinopathies but also for understanding the dynamics of seeding and spread of α-syn pathology. Previous observations in isolated REM sleep behaviour disorder showed that olfactory dysfunction can identify patients at high risk of short-term conversion to overt α-synucleinopathy (Mahlknecht et al., 2015). Intriguingly, in our study, α-syn RT-QuIC positivity in the OM from patients with isolated REM sleep behaviour disorder was preferentially associated with olfactory dysfunction, but this was not observed in Parkinson’s disease patients. A possible explanation could be that different olfactory areas are involved in the two conditions. For instance, olfactory dysfunction in Parkinson’s disease is driven by the involvement of the anterior olfactory nucleus and olfactory bulb or larger areas of the olfactory system beyond the OM, while isolated REM sleep behaviour disorder patients may have a more relevant involvement of peripheral sites such as OM. These differences might reflect rostral trans-synaptic propagation of α-syn pathology, as reported for the enteric neurons in Parkinson’s disease (Sprenger et al., 2015). The fact that the rate of α-syn RT-QuIC positivity in the OM of Parkinson’s disease patients was present in less than 50% of subjects might suggest that numbers of olfactory neurons progressively decrease with disease progression (Witt et al., 2009) a process which might occur also in isolated REM sleep behaviour disorder. To further investigate this hypothesis, prospective studies including isolated REM sleep behaviour disorder as well as early Parkinson’s disease subjects are needed. This approach is expected to provide further insight into the temporal dynamics of α-syn seeding activity in the OM and its association with phenoconversion. Future studies should include larger Parkinson’s disease samples in different disease stages to enable differential assessment of OM RT-QuIC in Parkinson’s disease subgroups based on in-depth phenotypic characterization.

In summary, we show that α-syn RT-QuIC enables detection of pathological seeding activity in olfactory neurons from patients with isolated REM sleep behaviour disorder and Parkinson’s disease. This reaction appears highly specific for these synucleinopathies as compared to controls and is associated with olfactory disturbances in isolated REM sleep behaviour disorder. Probing the OM with α-syn RT-QuIC may provide a valuable marker to recognize patients in an early-stage of α-synuclein related neurodegeneration and might help to select subjects for clinical disease-modification trials of interventions targeting α-syn pathologic conversion and spread.

## Data Availability

Data available upon request to the corresponding author

## Abbreviations list

α-syn: α-synuclein
IQR: Interquartile range
MDS: Movement disorders society
MDS-UPDRS: Movement disorders society Unified Parkinson’s Disease Rating Scale
MoCA: Montreal cognitive assessment
p-α-syn: Phosphorylated α-synuclein
OM: Olfactory mucosa
REM: Rapid eye movement
RBD1Q: REM sleep behaviour disorder single question
RT-QuIC: Real-Time Quaking Induced Conversion
SCOPA-AUT: SCales for Outcomes in PArkinson’s disease - AUTonomic dysfunction
ThT: Thioflavin T
UPSIT: University of Pennsylvania Smell Identification Test

## Acknowledgements

Authors deeply thank Santina Castriciano for donating nasal swabs.

## SUPPLEMENTARY TABLE

**Table S1.** Alpha-syn RT-QuIC assay, lag-phase, and final fluorescence values in OM from subjects with isolated RBD, Parkinson’s disease and healthy controls.

| **Positive RT-QuIC**<br>**in each subject group**<br>**(n/total)** | **Final fluorescence in**<br>**positive RT-QuIC (rfu)** | **Lag-phase in positive**<br>**RT-QuIC (hours)** |
| --- | --- | --- |
| **iRBD (28/63)** | 214720 ± 48471 | 47 ± 14.2 |
| **PD (19/41)** | 199415 ± 52560 | 45 ± 14 |
| **Controls (6/59)** | 194000 ± 38925 | 37 ± 12 |

Values of lag-phase and rfu are reported as mean ± standard deviations.

## References

American Academy of Sleep Medicine. The international classification of sleep disorders: diagnostic and coding manual. 3rd edition. Rev. ed. ed. Darien, Il.: American Academy of Sleep Med, 2014.

Antelmi E, Donadio V, Incensi A, Plazzi G, Liguori R. Skin nerve phosphorylated α-synuclein deposits in idiopathic REM sleep behavior disorder. Neurology 2017;88:2128–2131.

Arnold SE, Lee EB, Moberg PJ, et al. Olfactory epithelium amyloid-beta and paired helical filament-tau pathology in Alzheimer disease. Ann Neurol. 2010 67:462–9.

Beach TG, Adler CH, Lue L, et al. Unified staging system for Lewy body disorders: correlation with nigrostriatal degeneration, cognitive impairment and motor dysfunction. Acta Neuropathol 2009;117:613–34.

Bongianni M, Orrù C, Groveman BR, et al. Diagnosis of Human Prion Disease Using Real-Time Quaking-Induced Conversion Testing of Olfactory Mucosa and Cerebrospinal Fluid Samples. JAMA Neurol 2017;74:155–162.

Bongianni, M, Ladogana, A, Capaldi S et al. Alpha-synuclein RT-QuIC assay in CSF of patients with Dementia with Lewy bodies. Ann Clin Transl Neurol 2019:2120–2126.

Braak, Del Tredici K, Rüb U, de Vos RA, Jansen Steur EN, Braak E. Staging of brain pathology related to sporadic Parkinson’s disease. Neurobiol Aging 2003;24:197–211.

Brozzetti L, Sacchetto L, Cecchini MP, et al. Neurodegeneration-Associated Proteins in Human Olfactory Neurons Collected by Nasal Brushing. Frontiers in Neuroscience 2020,;14:145.

Chahine LM, Beach TG, Seedorff N, et al.; Systemic Synuclein Sampling study. Feasibility and Safety of Multicenter Tissue and Biofluid Sampling for α-Synuclein in Parkinson’s Disease: The Systemic Synuclein Sampling Study (S4). J Parkinsons Dis 2018;8:517–527.

De Luca CMG, Elia AE, Portaleone SM, et al. Efficient RT-QuIC seeding activity for α-synuclein in olfactory mucosa samples of patients with Parkinson’s disease and multiple system atrophy. Transl Neurodegener 2019;8:24.

Donadio V, Doppler K, Incensi A, et al. Abnormal α-synuclein deposits in skin nerves: intra- and inter-laboratory reproducibility. Eur J Neurol 2019;26:1245–1251.

Doppler K, Jentschke HM, Schulmeyer L, et al. Dermal phospho-alpha-synuclein deposits confirm REM sleep behaviour disorder as prodromal Parkinson’s disease. Acta Neuropathol 2017;133:535–545.

Doty RL. The smell identification test administration manual, 3rd edn. Philadephia: Sensonics, 1995.

Fairfoul G, McGuire LI, Pal S, et al. Alpha-synuclein RT-QuIC in the CSF of patients with alpha-synucleinopathies. Ann Clin Transl Neurol 2016;3:812–818.

Fernández-Arcos A, Vilaseca I, Aldecoa I, et al. Alpha-synuclein aggregates in the parotid gland of idiopathic REM sleep behavior disorder. Sleep Med 2018;52:14–17.

Garrido A, Fairfoul G, Tolosa ES, Martí MJ, Green A; Barcelona LRRK2 Study Group. α-synuclein RT-QuIC in cerebrospinal fluid of LRRK2-linked Parkinson’s disease. Ann Clin Transl Neurol 2019;6:1024–1032.

Gibb WR, Lees AJ. The relevance of the Lewy body to the pathogenesis of idiopathic Parkinson’s disease. J Neurol Neurosurg Neuropsychiatry 1988;51:745–752.

Goetz CG, Tilley BC, Shaftman SR, et al; Movement Disorder Society UPDRS Revision Task Force. Movement Disorder Society-sponsored revision of the Unified Parkinson’s Disease Rating Scale (MDS-UPDRS): scale presentation and clinimetric testing results. Mov Disord 2008;23:2129–2170.

Groveman BR, Orrù CD, Hughson AG, Raymond LD, Zanusso G, Ghetti B, Campbell KJ, Safar J, Galasko D, Caughey B. Rapid and ultra-sensitive quantitation of disease-associated α-synuclein seeds in brain and cerebrospinal fluid by αSyn RT-QuIC, Acta Neuropathol Commun 2018 9;6:7.

Heinzel S, Berg D, Gasser T, Chen H, Yao C, Postuma RB; MDS Task Force on the Definition of Parkinson’s Disease. Update of the MDS research criteria for prodromal Parkinson’s disease. Mov Disord 2019;34:1464–1470.

Högl B, Stefani A, Videnovic A. Idiopathic REM sleep behaviour disorder and neurodegeneration - an update. Nat Rev Neurol 2018;14:40–55.

Iranzo A, Tolosa E, Gelpi E, et al. Neurodegenerative disease status and postmortem pathology in idiopathic rapid-eye-movement sleep behaviour disorder: an observational cohort study. Lancet Neurol 2013;12:443–453.

Iranzo A, Borrego S, Vilaseca I, et al. α-Synuclein aggregates in labial salivary glands of idiopathic rapid eye movement sleep behavior disorder. Sleep 2018;41(8).

Lawton M, Hu MT, Baig F, et al. Equating scores of the University of Pennsylvania Smell Identification Test and Sniffin’ Sticks test in patients with Parkinson’s disease. Parkinsonism Relat Disord 2016;33:96–101.

Mahlknecht P, Iranzo A, Högl B, et al; Sleep Innsbruck Barcelona Group. Olfactory dysfunction predicts early transition to a Lewy body disease in idiopathic RBD. Neurology 2015;84:654–658.

Nasreddine ZS, Phillips NA, Bedirian V, et al. The Montreal Cognitive Assessment, MoCA: A brief screening tool for mild cognitive impairment. J Am Geriatr Soc 2005;53:695–699.

Oleszkiewicz A, Schriever VA, Croy I, Hähner A, Hummel T. Updated Sniffin’ Sticks normative data based in an extended sample of 9139 subjects. Eur Arch Otorhinolaryngol 2019;276:719–728.

Postuma RB, Arnulf I, Högl B, et al. A single-question screen for rapid eye movement sleep behavior disorder: a multicenter validation study. Mov Disord 2012;27:913–6.

Postuma RB, Berg D, Stern M, et al. MDS clinical diagnostic criteria for Parkinson’s disease. Mov Disord 2015;30:1591–1601.

Race B, Williams K, Chesebro B. Transmission studies of chronic wasting disease to transgenic mice overexpressing human prion protein using the RT-QuIC assay. Vet Res 2019;50:6.

Rey NL, Wesson DW, Brundin P. The olfactory bulb as the entry site for prion-like propagation in neurodegenerative diseases. Neurobiol Dis 2018;109:226–248.

Ruffmann C, Bengoa-Vergniory N, Poggiolini I, et al. Detection of alpha-synuclein conformational variants from gastro-intestinal biopsy tissue as a potential biomarker for Parkinson’s disease. Neuropathol Appl Neurobiol 2018;44:722–736.

Saito Y, Shioya A, Sano T, Sumikura H, Murata M, Murayama S. Lewy body pathology involves the olfactory cells in Parkinson’s disease and related disorders. Mov Disord 2016;31:135–8.

Schenck CH, Boeve BF, Mahowald MW. Delayed emergence of a parkinsonian disorder or dementia in 81% of older men initially diagnosed with idiopathic rapid eye movement sleep behavior disorder: a 16-year update on a previously reported series. Sleep Med 2013;14:744–748.

Sprenger FS, Stefanova N, Gelpi E, et al. Enteric nervous system α-synuclein immunoreactivity in idiopathic REM sleep behavior disorder. Neurology 2015;85:1761–1768.

van Rumund A, Green AJE, Fairfoul G, Esselink RAJ, Bloem BR, Verbeek MM. α-Synuclein real-time quaking-induced conversion in the cerebrospinal fluid of uncertain cases of parkinsonism. Ann Neurol 2019;85:777–781.

Vilas D, Iranzo A, Tolosa E, et al. Assessment of α-synuclein in submandibular glands of patients with idiopathic rapid-eye-movement sleep behaviour disorder: a case-control study. Lancet Neurol 2016;15:708–718.

Visser M, Marinus J, Stiggelbout AM, Van Hilten JJ. Assessment of autonomic dysfunction in Parkinson’s disease: the SCOPA-AUT. Mov Disord 2004;19:1306–1312.

Witt M, Bormann K, Gudziol V, et al. Biopsies of olfactory epithelium in patients with Parkinson’s disease. Mov Disord 2009;24:906–14.

